# Caries status, caries severity, and oral health-related quality of life of preschool children in Kuantan

**DOI:** 10.1101/2024.07.04.24309980

**Authors:** Muhammad Zaki Ramli, Nina Suhaity Azmi, Ahmad Faisal Ismail

## Abstract

Dental caries among preschool children is prevalent and may affect their oral health-related quality of life (OHRQOL). The aim of this study was to assess the oral health-related quality of life (OHRQOL) of preschool children from Kuantan in relation to their dental caries status and severity. A cross-sectional survey using the Malay version of Early Childhood Oral Health Impact Scale (Malay-ECOHIS) involving preschool children aged three until six years old was conducted employing a convenience sampling. The dmft score of the preschool children was evaluated by a professional pediatric dentist, and the body mass index (BMI) was recorded. The caries status data was classified into either Absent (dmft = 0) or Present (dmft ≥ 1), while caries severity data was categorized into No Caries (dmft = 0), Moderate (dmft = 1 or 2), and High (dmft ≥ 3), depending on the caries experience. The Pearson Chi-square and Spearman correlation analyses were conducted. A high caries prevalence was recorded (89.1%), affecting 55 preschool children. They were more impacted than their family in terms of their OHRQOL, and those with high caries severity and those from low household income showed the lowest OHRQOL. However, since most of them were from high household income, their families were not financially impacted. It is important to not only assess the caries status of the preschool children when assessing their OHRQOL, but also their caries severity.

## Introduction

Dental caries is the most prevalent disease in humans globally (1). It remains a significant public health problem and is considered pandemic worldwide. Untreated dental caries is one of the most prevalent noncommunicable diseases (2). A subset of dental caries, early childhood caries (ECC), is a considerable pediatric and public health problem worldwide (3–5). ECC is characterized by the presence of at least one decayed, missing, or filled tooth surfaces in any primary tooth, affecting children below 71 months old (6). A severe form of ECC, called severe early childhood caries (S-ECC) is an aggressive form of dental caries classified by the presence of a decayed, missing (due to caries), or filled tooth (dmft) index score of ≥4 (age 3), ≥5 (age 4), or ≥6 (age 5) (7).

Dental caries may significantly impact the quality of life of the individuals affected by the highly prevalent oral disease (8). Due to a poor oral health and practice, children’s quality of life can greatly be impacted (9). Distress could result following dental pain, dental abscess, gum disease and damaged teeth, diseases which may eventually lead to negative impacts on their social, functional, and psychological well-being (10). During these years of growth, children are actively attending schools, which are an effective platform for conducting oral health education programs (11).

However, since the end of 2019, the world was struck with the pandemic coronavirus (COVID-19) infection, thus the effects included the closure of schools, not allowing the school-aged children for schooling (12), including the preschool children. Not only the schools and preschools, but the dental services were also limited to be accessed during the period, thus opening more possibilities for the preschoolers to be affected by dental caries if they were not treated immediately. Also, not only the preschool children were affected; their whole family members could also be affected Hence, studies on this population’s oral health-related quality of life (OHRQOL) in relation to their dental caries status and severity need to be conducted. This was the aim of the study specifically on the preschool children from preschools in Kuantan, Pahang, Malaysia.

## Methodology

### Ethical Approval and Informed Consent

Ethical approval was granted by the IIUM Research Ethics Committee (IREC) (ID no.: IREC 2020-036). The data were collected from 1 September 2021 until 31 December 2021. Parents and children were informed that the participants could withdraw from the study at any time. A written consent was distributed to the parents before including their preschool children in the study.

### Data Collection

A cross-sectional survey involving preschool children aged three to six years old was conducted. A convenience sampling method was employed as few schools allowed outsiders to conduct studies at their premises during the pandemic. A validated questionnaire named Malay-Early Childhood Oral Health Impact Scale (Malay-ECOHIS) was distributed to the parents via the preschools. The inclusion criteria were those parents who can read and write in Malay language, and they must be a citizen of Malaysia. Parents of preschool children functioned as a proxy to answer the questionnaires. The questionnaire also collected information about the age and gender of the children and the family’s monthly household income.

The Malay-ECOHIS consisted of 13 items with two sections of the child impacts section and family impacts section. For answering this section, the parents were requested to recall the child’s all-inclusive lifetime experience. The 6-point Likert Scale was used as the response scale, namely “never”, “hardly ever”, “occasionally”, “often”, “very often,” and “don’t know.” Each of the scales was scored as 0 for “never”, 1 for “hardly ever”, 2 for “occasionally”, 3 for “often”, and 4 for “very often,” while “don’t know” responses were recorded as a missing value. Questionnaires with at least one “don’t know” answer on the child impacts section or family impacts section were excluded from being analyzed. The total score was calculated using a simple sum. The total score for the child impacts section was from 0 to 36, and the total score for family impacts section was from 0 to 16. To determine the presence of impact, the answer records at least one answer of “occasionally” or “often” or “very often,” while “never” or “hardly ever” indicated the absence of impact for both parts.

### Children’s Oral Examination

The dmft score of the preschool children was evaluated by a trained and calibrated pediatric dentist. Visual inspection was used for registration of dental caries, and the examinations were conducted in daylight using a plane mouth mirror.

### Measuring the Preschool Children’s Weight and Height

The body weight and height were recorded so that their body mass index (BMI) could be calculated. The BMI Percentile Calculator for Child and Teen by the Centers for Disease Control and Prevention (CDC) was used as the subjects in this study were preschool children. The calculator can be accessed at the following link: https://www.cdc.gov/healthyweight/bmi/calculator.html. The data was then classified into the BMI categories namely underweight, health, overweight, and obese.

### Data Analysis

Data processing, data analyses, and statistical evaluation were performed by means of the IBM SPSS Statistics 25. The prevalence proportion rates of dmft, BMI, and household income were calculated. The caries status data was classified into either Absent (dmft = 0) or Present (dmft ≥ 1), while caries severity data was categorized into No Caries (dmft = 0), Moderate (dmft = 1 or 2), and High (dmft ≥ 3), depending on the number of caries experience. For household income, the categories were Low, Moderate, and High based on Pahang’s 2019 mean household income as reported by the Department of Statistics Malaysia.

For the statistical analyses, to compare between groups, the Mann-Whitney U test was performed. Next, to compare the proportional means, univariant statistical analysis was performed using the Kruskal-Wallis non-parametric test. To assess the association and correlation between caries status and caries severity among preschool children with their oral health-related quality of life, the Pearson Chi-square and Spearman correlation analyses were conducted. The level of significance was set to p < 0.05.

## Results

Two preschools agreed to take part in the study and were visited to collect the data and samples from the preschool children. In total, 55 Malay preschool children participated in the study. Table 1 shows the sociodemographic characteristics of the preschool children from the two preschools, and Table 2 shows the summary of descriptive statistics of the preschool children. The majority of the preschool children had caries (89.1%) with majority of them had high severity (65.5%), were of healthy BMI (47.3%), and came from families with high household income (52.7%). Only household income showed a significant association with caries severity, while no significant association was found with caries status (Table 3).

**Table 1.** Sociodemographic characteristics of the preschool children from the two preschools.

| Sociodemographic characteristics | Frequency |  |
| --- | --- | --- |
|  | n | % |
| **Age (years)** |  |  |
| 3 | 5 | 9.1 |
| 4 | 18 | 32.7 |
| 5 | 15 | 27.3 |
| 6 | 17 | 30.9 |
| **Gender** |  |  |
| Boys | 32 | 58.2 |
| Girls | 23 | 41.8 |
| **Household income category** |  |  |
| High | 29 | 52.7 |
| Middle | 20 | 36.4 |
| Low | 6 | 10.9 |
| **BMI (type)** |  |  |
| Underweight | 11 | 20.0 |
| Healthy | 26 | 47.3 |
| Overweight | 8 | 14.5 |
| Obese | 10 | 18.2 |
| **Dental caries status** |  |  |
| Present (dmft ≥ 1) | 49 | 89.1 |
| Absent (dmft = 0) | 6 | 10.9 |
| **Dental caries severity** |  |  |
| Caries free (dmft = 0) | 6 | 10.9 |
| Moderate (dmft = 1 or 2) | 13 | 23.6 |
| High (dmft ≥ 3) | 36 | 65.5 |

**Table 2.** Descriptive statistics.

| **N=55** | **Mean** | **Range** |
| --- | --- | --- |
| Age | 4.8 | 3–6 |
| Weight (kg) | 18.2 | 12.9–34.9 |
| Height (cm) | 102.26 | 91–121 |
| BMI | 16.5 | 13.43–17.54 |
| dmft | 4.47 | 0–14 |
| Household income (RM) | 7,751.11 | 1,000–30,000 |

**Table 3.** Association between caries status, severity, and household income categories.

|  | **Household Income Categories** |  |  | **p-value\*** |
| --- | --- | --- | --- | --- |
|  | **Low**<br>**N (%)** | **Middle**<br>**N (%)** | **High**<br>**N (%)** |  |
| **Caries status** |  |  |  |  |
| Absent | 0 (0.0%) | 4 (7.3%) | 2 (3.6%) | .23<sup>(a)</sup> |
| Present | 6 (10.9%) | 16 (29.1%) | 27 (49.1%) |  |
| **Caries severity** |  |  |  |  |
| Absent | 0 (0.0%) | 4 (7.3%) | 2 (3.6%) | .05<sup>(b)</sup> |
| Moderate | 0 (0.0%) | 2 (3.6%) | 11 (20.0%) |  |
| High | 6 (10.9%) | 14 (25.5%) | 16 (29.1%) |  |

\* Chi-square test; (a) $\chi^2$ (2, N = 55) = 2.916; (b) $\chi^2$ (4, N = 55) = 9.722

Table 4 shows the Malay-ECOHIS responses of parents (n=55) about the oral conditions of their children. In the child impacts section, more than half of the children were reported to never have “had pain in the teeth, mouth or jaws” (56.4%). Next, the most frequently reported oral impact was “difficulty eating some food” with 14.5% of the children were reported to hardly ever experience the difficulty. Only 29.1% of the children were reported to experience the oral impact namely difficulty to eat some food, while the rest of the children (70.9%) were reported to never experience it. As for the other oral impacts to the children, most of the children were reported to never experience any of these, ranging from 85.5% to 92.7% of the children. However, it is notable that six (10.9%) children were reported to occasionally avoid smiling or laughing when they were orally impacted.

**Table 4.** Malay-ECOHIS responses of parents (n=55)

| Impacts | Never |  | Hardly ever |  | Occasionally |  | Often |  | Very often |  | Don't know |  | Mean (SD) |
| --- | --- | --- | --- | --- | --- | --- | --- | --- | --- | --- | --- | --- | --- |
|  | n | % | n | % | n | % | n | % | n | % | n | % |  |
| **Child Impacts Section** |  |  |  |  |  |  |  |  |  |  |  |  |  |
| Child Symptom Domain |  |  |  |  |  |  |  |  |  |  |  |  |  |
| How often has your child had pain in the teeth, mouth or jaws | 31 | 56.4 | 15 | 27.3 | 8 | 14.5 | 0 | 0.0 | 1 | 1.8 | 0 | 0.0 | 0.64 (0.87) |
| Child Function Domain |  |  |  |  |  |  |  |  |  |  |  |  |  |
| How often has your child ..... because of dental problems or dental treatments? |  |  |  |  |  |  |  |  |  |  |  |  |  |
| had difficulty drinking hot or cold beverages | 50 | 90.9 | 3 | 5.5 | 2 | 3.6 | 0 | 0.0 | 0 | 0.0 | 0 | 0.0 | 0.13 (0.43) |
| had difficulty eating some food | 39 | 70.9 | 8 | 14.5 | 7 | 12.7 | 1 | 1.8 | 0 | 0.0 | 0 | 0.0 | 0.45 (0.79) |
| had difficulty pronouncing any words | 48 | 87.3 | 1 | 1.8 | 3 | 5.5 | 1 | 1.8 | 0 | 0.0 | 2 | 3.6 | 0.36 (1.09) |
| missed preschool, day care or school | 51 | 92.7 | 3 | 5.5 | 1 | 1.8 | 0 | 0.0 | 0 | 0.0 | 0 | 0.0 | 0.09 (0.35) |
| Child Psychology Domain |  |  |  |  |  |  |  |  |  |  |  |  |  |
| had trouble sleeping | 49 | 89.1 | 4 | 7.3 | 2 | 3.6 | 0 | 0.0 | 0 | 0.0 | 0 | 0.0 | 0.15 (0.45) |
| been irritable or frustrated | 49 | 89.1 | 3 | 5.5 | 3 | 5.5 | 0 | 0.0 | 0 | 0.0 | 0 | 0.0 | 0.16 (0.50) |
| Child Self-Image and Social Interaction Domain |  |  |  |  |  |  |  |  |  |  |  |  |  |
| avoided smiling or laughing | 47 | 85.5 | 2 | 3.6 | 6 | 10.9 | 0 | 0.0 | 0 | 0.0 | 0 | 0.0 | 0.25 (0.64) |
| avoided talking | 49 | 89.1 | 2 | 3.6 | 5 | 9.1 | 0 | 0.0 | 0 | 0.0 | 1 | 1.8 | 0.18 (0.55) |
| **Family Impacts Section** |  |  |  |  |  |  |  |  |  |  |  |  |  |
| Parental Distress Domain |  |  |  |  |  |  |  |  |  |  |  |  |  |
| How often have you or another family member<br>..... because of your child's dental<br>problems or treatment? |  |  |  |  |  |  |  |  |  |  |  |  |  |
| been upset | 45 | 81.8 | 2 | 3.6 | 4 | 7.3 | 0 | 0.0 | 0 | 0.0 | 0 | 0.0 | 0.27 (0.68) |
| felt guilty | 36 | 65.5 | 10 | 18.2 | 3 | 5.5 | 4 | 7.3 | 0 | 0.0 | 2 | 3.6 | 0.69 (1.23) |
| Family Function Domain |  |  |  |  |  |  |  |  |  |  |  |  |  |
| taken time off from work | 54 | 98.2 | 1 | 1.8 | 0 | 0.0 | 0 | 0.0 | 0 | 0.0 | 0 | 0.0 | 0.02 (0.13) |
| How often has your child had dental problems or dental treatments that had a financial impact on your family? | 53 | 96.4 | 0 | 0.0 | 0 | 0.0 | 0 | 0.0 | 0 | 0.0 | 2 | 3.6 | 0.18 (0.94) |

Next, in terms of family impacts, feeling guilty by the parent or another family member because the children had dental problem or treatment was reported to score the lowest among the items. The other two items under the family impacts showed different distributions. The item “been upset” was mostly rated as “Never” by 45 (81.8%) of the parents. The final item in the family impacts, “taken time off from work” was rated by almost all parents, 54 (98.2%) as “Never”. Finally, the last item in the Malay-ECOHIS asked the parents about how often their children had dental problems or dental treatments that had a financial impact on their family. From the survey, majority of the parents (96.4%) rated that they never had such an issue. In this study, the Malay-ECOHIS scores ranged from 0 to 15 in the child impacts section (mean = 2.4, SD = 3.5), while for the family impacts section, the scores ranged from 0 to 8 (mean = 1.2, SD = 2.0).

Next, the data collected from the parents was classified into either having no impact on the children and family or not. This was conducted by recording “never” or “hardly ever” as the absence of impact, while “occasionally,” “often,” and “very often” were grouped into the presence of impact. This section disregarded those with at least one “don’t know” answer on the child impacts section or family impacts section. Of the 55 respondents, 4 included “don’t know” in at least of the answers, thus they were excluded in the analysis, leaving 51 respondents for further analyses.

As shown in Table 5, in both sections and domains, the “never and hardly ever” responses recorded much higher scores than their counterparts namely “occasionally, often, and very often”. However, it can be noted that the child symptom domain had the highest scores for the “occasionally, often, and very often” responses, followed by “had difficulty eating some food” item under the child function domain and “felt guilty” item under the parental distress domain, which is in the family impacts section. Moreover, in the family impacts section, no responses were recorded for “occasionally, often, and very often” specifically from the “taken time off from work” item under the family function domain, as well as the financial impact item.

**Table 5.** Malay-ECOHIS responses of parents (n=51)

| Impacts | Never or hardly ever |  | Occasionally, often, or very often |  |
| --- | --- | --- | --- | --- |
|  | n | % | n | % |
| **Child Impacts Section** |  |  |  |  |
| Child Symptom Domain |  |  |  |  |
| How often has your child had pain in the teeth, mouth or jaws | 43 | 84.3 | 8 | 15.7 |
| Child Function Domain |  |  |  |  |
| How often has your child ..... because of dental problems or dental treatments? |  |  |  |  |
| had difficulty drinking hot or cold beverages | 49 | 96.1 | 2 | 3.9 |
| had difficulty eating some food | 44 | 86.3 | 7 | 13.7 |
| had difficulty pronouncing any words | 48 | 94.1 | 3 | 5.9 |
| missed preschool, day care or school | 50 | 98.0 | 1 | 2.0 |
| Child Psychology Domain |  |  |  |  |
| had trouble sleeping | 49 | 96.1 | 2 | 3.9 |
| been irritable or frustrated | 49 | 96.1 | 2 | 3.9 |
| Child Self-Image and Social Interaction Domain |  |  |  |  |
| avoided smiling or laughing | 46 | 90.2 | 5 | 9.8 |
| avoided talking | 49 | 96.1 | 2 | 3.9 |
| **Family Impacts Section** |  |  |  |  |
| Parental Distress Domain |  |  |  |  |
| How often have you or another family member .....<br>because of your child's dental problems or treatment? |  |  |  |  |
| been upset | 49 | 96.1 | 2 | 3.9 |
| felt guilty | 45 | 88.2 | 6 | 11.8 |
| Family Function Domain |  |  |  |  |
| taken time off from work | 51 | 100.0 | 0 | 0.0 |
| How often has your child had dental problems or dental treatments that<br>had a financial impact on your family? | 51 | 100.0 | 0 | 0.0 |

Next, the association and correlation between caries status and children’s OHRQOL and family’s OHRQOL in the Malay-ECOHIS among the preschool children were assessed. As shown in Table 6, between the impacts on the children and the family, the score was higher on the children compared to the family. This was evident in the score for impacts on the children with caries (25.5%) than on the family members with caries-bearing children (11.8%).

**Table 6.**
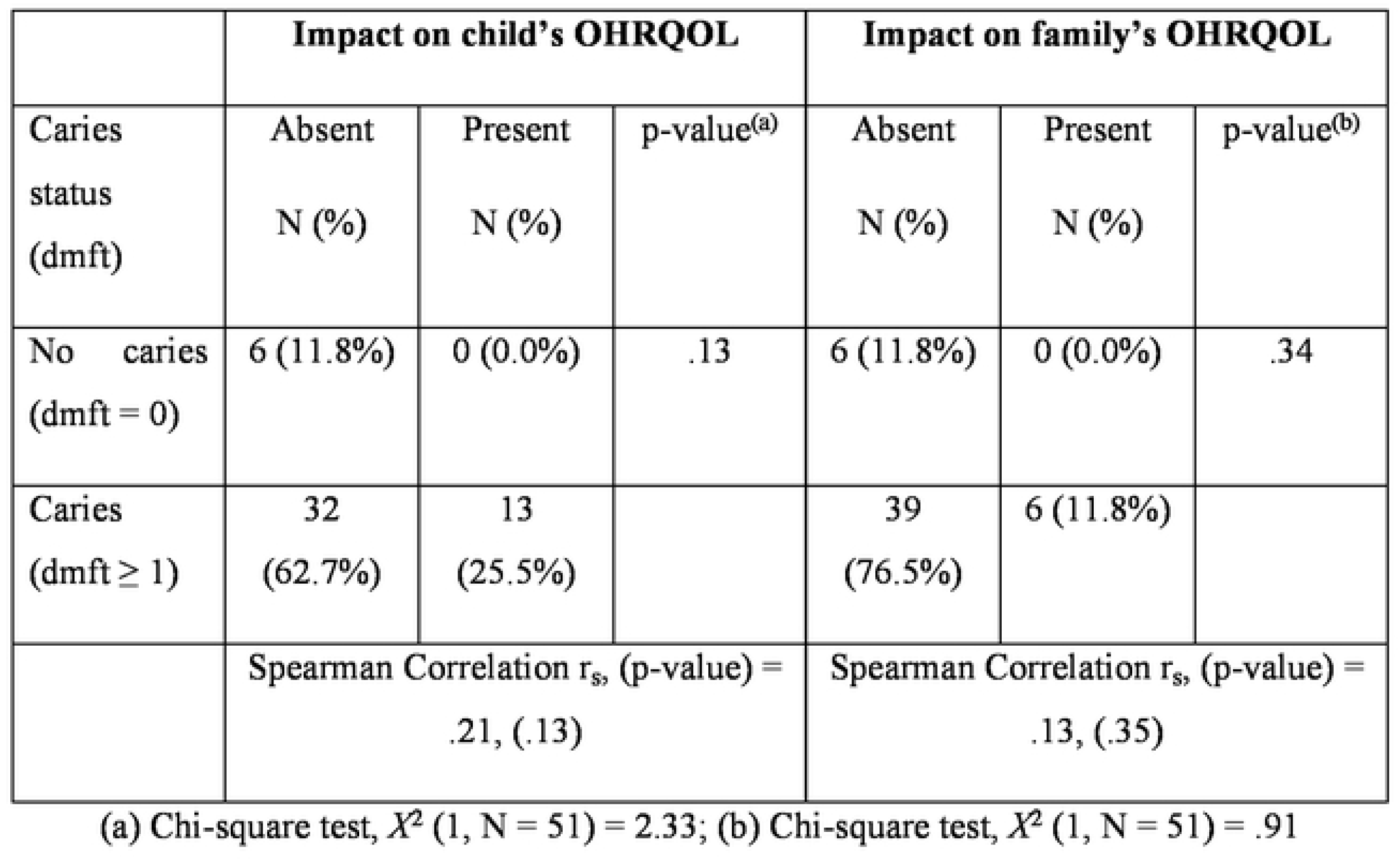
Association and correlation between caries status and child’s OHRQOL and family’s OHRQOL in the Malay-ECOHIS.

However, both did not show any significant correlation (p>0.05) as also evidenced by the Spearman correlation results.

As shown in Table 7, none of the domains and sections had a p-value of less than 0.05. Therefore, caries status did not have a significant impact on any of the sections and domains in the Malay-ECOHIS. Also notable are the means for the preschool children with caries scored higher than the means for the preschool children without caries in all sections and domains. Thus, the caries-bearing children were more impacted in terms of their OHRQOL compared to the caries-free children. The table also shows that high caries severity had the highest means in all sections and domains, as well as the Malay-ECOHIS score. Both child impacts and family impacts sections had statistically significant difference (p < .05), but not in all domains. Overall, the Malay-ECOHIS showed the highest mean in the high caries severity group.

**Table 7.** Descriptive statistics of the Malay-ECOHIS responses, ranges, and comparisons according to their caries status and caries severity (n=51)

| Variables | Items (N) | Possible Range | Mean (SD) | Caries Status |  | p-value* | Caries Severity |  |  | p-value* | Pairwise p-value |
| --- | --- | --- | --- | --- | --- | --- | --- | --- | --- | --- | --- |
|  |  |  |  | Absent | Present |  | No caries<sup>1</sup> | Moderate<sup>2</sup> | High<sup>3</sup> |  |  |
| **Child Impacts Section** | 9 | 0–36 | 11.06 (3.32) | 9.83 (1.60) | 11.22 (3.46) | .40 | 9.83 (1.60) | 9.15 (0.38) | 12.06 (3.80) | **.03** | **.00**<br>(2 vs. 3) |
| Child symptom domain | 1 | 0–4 | 1.63 (0.87) | 1.33 (0.52) | 1.67 (0.90) | .52 | 1.33 (0.52) | 1.15 (0.38) | 1.88 (0.98) | **.02** | **.02**<br>(2 vs. 3) |
| Child function domain | 4 | 0–16 | 4.80 (1.61) | 4.33 (0.82) | 4.87 (1.69) | .58 | 4.33 (0.82) | 4.00 (0.00) | 5.22 (1.90) | **.01** | **.01**<br>(2 vs. 3) |
| Child psychology domain | 2 | 0–8 | 2.27 (0.70) | 2.17 (0.41) | 2.29 (0.73) | 1.00 | 2.17 (0.41) | 2.00 (0.00) | 2.41 (0.84) | .19 | - |
| Child self-image and social interaction domain | 2 | 0–8 | 2.35<br>(0.98) | 2.00<br>(0.00) | 2.40<br>(1.03) | .56 | 2.00<br>(0.00) | 2.00 (0.00) | 2.56<br>(1.19) | .10 | - |
| **Family impacts section** | 4 | 0–16 | 4.75<br>(1.37) | 4.12<br>(0.41) | 4.82<br>(1.43) | .43 | 4.17<br>(0.41) | 4.00 (0.00) | 5.16<br>(1.59) | **.01** | **.01**<br>**(2 vs. 3)** |
| Parental distress domain | 2 | 0–8 | 2.73<br>(1.34) | 2.17<br>(0.41) | 2.80<br>(1.41) | .43 | 2.17<br>(0.41) | 2.00 (0.00) | 3.12<br>(1.56) | **.01** | **.01**<br>**(2 vs. 3)** |
| Family function domain | 2 | 0–8 | 2.02<br>(0.14) | 2.00<br>(0.00) | 2.02<br>(0.15) | .94 | 2.00<br>(0.00) | 2.00 (0.00) | 2.03<br>(0.18) | .74 | - |
| Malay-ECOHIS<br>items | 13 | 0–52 | 15.80<br>(4.49) | 14.00<br>(2.00) | 16.04<br>(4.69) | .35 | 14.00<br>(2.00) | 13.15<br>(0.38) | 17.22<br>(5.12) | **.00** | **.02**<br>**(2 vs. 3)** |

\* Chi-square test

Finally, in Table 8, it shows that the family impacts section and both domains in the section had a statistically significant difference, with all of them had a statistically significant difference between high household income and low household income, while the family function domain also had a statistically significant difference between middle household income and low household income. As for the child impacts section, all except one domain did not have a significant difference. The child self-image and social interaction domain had a significant difference between two pairs, namely middle household income vs. low household income and high household income vs. low household income. In all sections and domains in the Malay-ECOHIS responses, the household income in the low category showed the highest means. The overall Malay-ECOHIS thus also scored the highest mean in the low household income.

**Table 8.** Descriptive statistics of the Malay-ECOHIS responses, ranges, and comparisons according to their household income (n=51)

| Variables | Items (N) | Possible Range | Mean (SD) | Household Income |  |  | p-value* | Pairwise p-value |
| --- | --- | --- | --- | --- | --- | --- | --- | --- |
|  |  |  |  | Low<sup>1</sup> | Middle<sup>2</sup> | High<sup>3</sup> |  |  |
| **Child impacts section** | 9 | 0–36 | 11.06 (3.32) | 14.33 (5.43) | 10.65 (2.66) | 10.60 (2.86) | .08 | - |
| Child symptom domain | 1 | 0–4 | 1.63 (0.87) | 2.33 (0.82) | 1.55 (0.76) | 1.52 (0.92) | .05 | - |
| Child function domain | 4 | 0–16 | 4.80 (1.61) | 5.50 (2.74) | 4.75 (1.33) | 4.68 (1.52) | .53 | - |
| Child psychology domain | 2 | 0–8 | 2.27 (0.70) | 2.83 (1.33) | 2.25 (0.64) | 2.16 (0.47) | .33 | - |
| Child self-image and social interaction domain | 2 | 0–8 | 2.35 (0.98) | 3.67 (1.97) | 2.10 (0.45) | 2.24 (0.72) | .01 | .01 (2 vs. 1)<br>.03 (3 vs. 1) |
| **Family impacts section** | 4 | 0–16 | 4.75 (1.37) | 6.83 (2.48) | 4.50 (0.83) | 4.44 (0.92) | .02 | .02 (3 vs. 1) |
| Parental distress domain | 2 | 0–8 | 2.73 (1.34) | 4.67 (2.50) | 2.50 (0.83) | 2.44 (0.92) | .03 | .03 (3 vs. 1) |
| Family function domain | 2 | 0–8 | 2.02 (0.14) | 2.17 (0.41) | 2.00 (0.00) | 2.00 (0.00) | .02 | .03 (2 vs. 1)<br>.03 (3 vs. 1) |
| Malay-ECOHIS items | 13 | 0–52 | 15.80 (4.49) | 21.17 (7.63) | 15.15 (3.23) | 15.04 (3.68) | .06 | - |

\* Chi-square test

## Discussion

This present study was the first to assess the impacts of demographic profiles, caries status, caries severity, BMI categories, and household income on the OHRQOL of the population aged three to six years old in Kuantan by adopting the Malay version of ECOHIS. This allowed the findings to be compared with the previous studies that used the same scale either in Malay, English or other languages. Also, by adopting an instrument that has been validated in the same country, it allows differences in semantics and norms to be factored out (13).

All the subjects in this study were Malays. In terms of ethnicity, this present study was equivalent to a previous study that involved an all-Malay population in Kota Bharu, Kelantan (14). Other earlier studies also did not manage to involve many preschool children from other ethnicities in Malaysia. For example, of the 138 subjects in the study by (15), only 8.7% were non-Malays, while only 2.4% of the 127 preschool children were non-Malays in the study by (16). Therefore, it was common to include mostly Malay subjects, if not all, in the study that used the Malay-ECOHIS questionnaire. Furthermore, among the preschool children, dental caries was present in most of them (89.1%), and the mean dmft score was 4.47 (range 0–14). This finding was in fact higher than the previous national oral health report of preschool children in Pahang (17) and a recent study from the similar population in Kuantan, Pahang (18).

Moreover, in terms of age, this study managed to collect responses from the parents and guardians of preschool children aged three to six years old. Most of the preschool children were four years old (32.7%). The three years old preschool children (9.1%) were the lowest in number among the subjects as usually in Malaysia these young children are not sent to preschools but instead are taken care off by their parents or grandparents. Some parents also did not allow their young children of that age to be involved in the study, since the sampling period was conducted during the pandemic.

Next, this present study found that caries was more prevalent in male preschool children (52.7%) compared to their female counterparts (36.4%) (Table 4). This finding was same as the one reported by previous studies (14,19). However, like that reported by (14), no significant association was established between the caries experience and gender, while (19) reported a significant association. However, earlier studies have reported gender as a risk factor for caries in preschool children, with being male to be a sociodemographic factor that was more frequently implicated compared to the female counterpart (20). Gender did not exhibit a direct significant relationship towards dmft, but it showed an indirect significant relationship towards dmft through children sociological factor (21).

None of the variables included in the study showed significant associations with either caries status or caries severity, except the association between household income categories and caries severity (p = .05). The high household income group with high caries severity scored the highest percentage (Table 3). However, the socioeconomic bias of this study should not be neglected. The preschool children were schooled at private schools that normally charge higher fees than the public preschools. Thus, it could be expected that the families were from high socioeconomic groups, thus had high household income. This finding was in congruence with (22). They stated children whose parents belong to an upper socioeconomic class were at higher odds of suffering from S-ECC. On the other hand, another study reported that the prevalence of S-ECC significantly decreased in children from high-income families (23). However, there are other factors such as the preschool children’s dietary intakes and accessibility to dental services that determine the association between caries severity and socioeconomic status of the preschool children’s families. Thus, these need to be further investigated.

From the distribution of items in this present study, among the 51 responses (Table 5), it was found that all the items in the Malay-ECOHIS never or hardly ever impacted the preschool children at much higher percentages than them being occasionally, often, and very often impacted. However, the “pain in the teeth, mouth or jaws” and “had difficulty eating some food” items in the child impacts section and the “felt guilty” item in the family impacts section were the three most frequently impacted items. Since the prevalence of caries in population of the present study was high, it could be expected that they scored highest in the child symptom domain as individuals with caries are reported to be more likely to have dental pain (24).

Other than that, severe caries can cause dental pain that exerts a negative impact on the performance of activities of daily living including going to school (25). However, in terms of the item with the least impact particularly in the child impacts section, this study found that “missed school” scored the least negative impact on the preschool children (Table 5). This finding was consistent with A. N. Hashim et al. (2015), Dolah et al. (2019), Montoya et al. (2021), and Arrow et al. (2021) as they also scored the lowest impact in the item. This means the least negative impact due to oral health issues among the children was their absenteeism from the learning institution. However, because the study was conducted during the pandemic, it could explain the reasons this item scored the least because the schools were closed for some months along the period.

Moreover, in the present study’s family impacts section of the Malay-ECOHIS, the “felt guilty” item scored the highest score among the items (Table 5). This was also in agreement with the earlier Malaysian studies using the same instrument, namely A. N. Hashim et al. (2015) and Dolah et al. (2019). It was reported that the number of parents who felt guilty increased with the increase of their children’s dental caries severity (27). Parents could feel guilty about the oral problems faced by their children as these may affect the quality of life of the children. The feeling of guilt was caused by the sense of responsibility among the parents for their children’s oral health problems (14). Despite the efforts made by the dental team in promoting the ways to prevent oral diseases, the parents have failed to take necessary and effective means to help their children maintain good oral practices as not to be affected by the eventual oral problems.

Financial difficulty could lead to a high prevalence of decayed teeth as they remain untreated (28), thus leading to the consequent oral impacts and eventually affecting the quality of life of the impacted individuals. However, this was not the case for the present study as the financial impact item scored among the lowest. This was possibly because the preschool children were mostly from high socioeconomic status families, thus they could afford to send their kids to private preschools. People from this subpopulation are also expected to be financially capable of getting dental healthcare and services, thus they are not substantially affected when it comes to their OHRQOL. Hence, this could also explain the reasons the parents and guardians involved in the study were not financially burdened.

When compared between the child impacts section and the family impacts section (Table 6), this present study found that the preschool children were more impacted than their family members, and neither had an association and a correlation with caries status of the children. This finding was inconsistent with the previous studies in a few ways. For example, Dolah et al. (2019) reported that the family members were more impacted than the children, and the association between the impacts on the family members and caries status was found to be significant (p < .05). Next, although A. N. Hashim et al. (2015) and Randrianarivony et al. (2020) also reported that the children were more impacted than their family members, they found that both had significant associations with caries status. These variations in the results were possibly contributed by the different demographic settings of the present study and the previous ones, besides the socioeconomic bias this present study had and due to the sample size variations.

As stated above, there was no association and correlation between the child impacts section and caries status. When these were further investigated at domain levels, none of the domains showed a significant difference with the caries status. The same findings were also found in the family impacts section. However, when the caries status was further divided into its severity levels, some significant differences were found. These include the significant differences between moderate and high caries severity in both sections and the following domains: child symptom domain, child function domain, and parental distress domain. In fact, the overall Malay-ECOHIS showed a significant difference between the moderate and high caries severity. Therefore, it is important to not only assess the caries status of the preschool children when assessing their OHRQOL, but also their caries severity as these two variables might give different results in certain sections and domains.

Moreover, in terms of the household income categories, the family impacts section and both of its domains recorded significant differences between low and high household income categories. Besides, a domain in the child impacts section, namely child self-image and social interaction domain also showed a significant difference between the same household income categories. More importantly, the low household income category showed the worst impacts in all sections and domains of the Malay-ECOHIS. These findings were expected because families from low socioeconomic backgrounds faced difficulties to keep their children’s dental healthcare due to factors such as financial reasons and access to dental care (29,30).

The study initially targeted to get the number of samples similar to the study by (14), i.e., around 200 preschool children. The study was conducted in a similar area in the east coast of Peninsular Malaysia, i.e., Kota Bharu, Kelantan. However, the present study managed to collect 55 samples during the pandemic. However, in studies on psychometric analysis, their sample size requirement is dependent mainly on the number of items in the scale. According to quality criteria for measurement properties of health status questionnaires, at least 50 subjects are necessary for an appropriate analysis of construct validity, reproducibility, and responsiveness (31). Hence, as the Malay-ECOHIS instrument used in this study contained 13 items, the sample size of 55 was considered sufficient for the purpose of assessing its psychometric properties. The current study was conducted during the pandemic at which it was difficult to arrange for data and sample collection with the preschools due to the changes in the rules set by the Malaysian government from time to time to control the coronavirus (COVID-19) infection. Also, some parents were reluctant to participate and allow their children to be involved in the study.

## Conclusion

Most of the preschool children (89.1%) in this study had caries (n=55), and the oral disease had impacted their quality of life. However, the preschool children were more impacted than their family. Other than that, their family was also affected, especially in feeling guilty. Since most of them came from families with a high household income background, they were not financially impacted. Caries status did not significantly affect these schools’ preschool children and their families during the pandemic in terms of their OHRQOL, but those with high caries severity and those from low household income significantly showed higher impacts. When assessing their OHRQOL, it is important to not only assess the caries status of the preschool children, but also their caries severity. Parents should take precautionary measures and actions as soon as possible to ensure their children are not affected by low OHRQOL at their later ages.

## Data Availability

The data underlying the results presented in the study are available from the authors. Please contact for the details.

## Acknowledgements

We would like to thank Glycobio for the supports and Universiti Malaysia Pahang for the grant (PGRS2003125).

## Author Contributions

Conceptualization: Muhammad Zaki Ramli

Data curation: Muhammad Zaki Ramli

Formal analysis: Muhammad Zaki Ramli

Funding acquisition: Muhammad Zaki Ramli, Nina Suhaity Azmi

Investigation: Muhammad Zaki Ramli

Methodology: Muhammad Zaki Ramli, Ahmad Faisal Ismail

Project administration: Muhammad Zaki Ramli

Supervision: Nina Suhaity Azmi, Ahmad Faisal Ismail

Validation: Nina Suhaity Azmi, Ahmad Faisal Ismail

Writing – original draft: Muhammad Zaki Ramli

Writing – review & editing: Muhammad Zaki Ramli, Nina Suhaity Azmi, Ahmad Faisal Ismail

## References

1. Prados-Privado M, Villalón JG, Martínez-Martínez CH, Ivorra C, Prados-Frutos JC. Dental caries diagnosis and detection using neural networks: A systematic review. J Clin Med. 2020;9(11):1–13.

2. Arantes R, Welch JR, Tavares FG, Ferreira AA, Jr CEAC, Vettore MV. Human ecological and social determinants of dental caries among the Xavante Indigenous people in Central Brazil. PLoS One. 2018;13(12):1–20.

3. Nagarajappa R, Satyarup D, Naik D, Dalai RP. Feeding practices and early childhood caries among preschool children of Bhubaneswar, India. European Archives of Paediatric Dentistry. 2020;21(1):67–74.

4. Folayan MO, Oginni AB, El Tantawi M, Alade M, Adeniyi AA, Finlayson TL. Association between nutritional status and early childhood caries risk profile in a suburban Nigeria community. Int J Paediatr Dent. 2020;30(6):798–804.

5. Li J, Fan W, Zhou Y, Wu L, Liu W, Huang S. The status and associated factors of early childhood caries among 3-to 5-year-old children in Guangdong, Southern China: A provincial cross-sectional survey. BMC Oral Health. 2020;20(1):1–8.

6. Anil S, Anand PS. Early childhood caries: Prevalence, risk factors, and prevention. Frontiers in Pediatrics. 2017;5(July):1–7.

7. Policy on Early Childhood Caries (ECC): Classifications, Consequences, and Preventive Strategies. American Academy of Pediatric Dentistry. 2016;(6):60–2.

8. Wang Y, Wang S, Wu C, Chen X, Duan Z, Xu Q, et al. Oral microbiome alterations associated with early childhood caries highlight the importance of carbohydrate metabolic activities. American Society for Microbiology. 2019;4(6):1–15.

9. Barasuol JC, Santos PS, Moccelini BS, Magno MB, Bolan M, Martins-Júnior PA, et al. Association between dental pain and oral health-related quality of life in children and adolescents: A systematic review and meta-analysis. Community Dentistry and Oral Epidemiology. 2020;48(4):257–63.

10. Aishah E, Nordin B, Shoaib LA, Yuzadi Z, Yusof M, Manan NM. Oral health-related quality of life among 11–12 year old indigenous children in Malaysia. BMC Oral Health. 2019;19(152).

11. Bramantoro T, Santoso CMA, Hariyani N, Setyowati D, Zulfiana AA, Nor NAM, et al. Effectiveness of the school-based oral health promotion programmes from preschool to high school: A systematic review. PLoS One. 2021;16(8 August):1–16.

12. Zierer K. Effects of pandemic-related school closures on pupils’ performance and learning in selected countries: A rapid review. Education Sciences. 2021;11(6).

13. Alwattban RR, Alkhudhayr LS, Al-Haj Ali SN, Farah RI. Oral health-related quality-of-life according to dental caries severity, body mass index and sociodemographic indicators in children with special health care needs. Journal of Clinical Medicine. 2021;10(21):1–13.

14. Dolah S, Eusufzai SZ, Alam MK, Wan Ahmad WMA. Factors influencing oral health-related quality of life among preschool children in district of Kota Bharu, Malaysia: A cross-sectional study. Pesqui Bras Odontopediatria Clin Integr. 2019;20:1–10.

15. Hashim NA, Mohd Yusof ZY, Saud R. Responsiveness to change of the Malay-ECOHIS following treatment of early childhood caries under general anaesthesia. Community Dent Oral Epidemiol. 2018;1–8.

16. Hashim AN, Yusof ZYM, Esa R. The Malay version of the Early Childhood Oral Health Impact Scale (Malay-ECOHIS) - assessing validity and reliability. Health Qual Life Outcomes. 2015;13(1):1–10.

17. National Oral Health Survey of Preschool Children 2015 (NOHPS 2015). 2015.

18. Ismail AF, Adon AA, Nur A, Husain F, Sukmasari S, Ardini YD. Association between caries experience and body mass index (BMI) among preschool children in Kuantan. Eurasian J Biosci. 2020;14:4363–6.

19. Duangthip D, Chen KJ, Gao SS, Lo ECM, Chu CH. Early childhood caries among 3-to 5-year-old children in Hong Kong. International Dental Journal. 2019;69(3):230–6.

20. Kirthiga M, Murugan M, Saikia A, Kirubakaran R. Risk factors for early childhood caries: A systematic review and meta-analysis of case control and cohort studies. Pediatric dentistry. 2019;41(2):95–112.

21. Ismail NS, Abdul Ghani NM, Supaat S, Kharuddin AF, Ardini YD. The early childhood oral health impact scale (ECOHIS): Assessment tool in oral health related quality of life. Journal of International Dental and Medical Research. 2018;11(1):162–8.

22. Mansoori S, Mehta A, Ansari MI. Factors associated with oral health-related quality of life of children with severe early childhood caries. Journal of Oral Biology and Craniofacial Research. 2019;9(3):222–5.

23. Li Y, Wulaerhan J, Liu Y, Abudureyimu A, Zhao J. Prevalence of severe early childhood caries and associated socioeconomic and behavioral factors in Xinjiang, China: A cross-sectional study. BMC Oral Health. 2017;17(1):1–10.

24. Montoya ALB, Knorst JK, Uribe IMP, González RAB, Ardenghi TM, Sánchez CCA. Cross-cultural adaptation and psychometric properties of the Mexican version of the Early Childhood Oral Health Impact Scale (ECOHIS). Health and Quality of Life Outcomes. 2021;19(1):1–8.

25. Utami SP, Liza F, Lisfrizal H. The development of the impact of early childhood caries on the quality of life of children aged 3-5 years at Paedodonti RSGM Baiturrahmah. Denta. 2021;15(2):92–9.

26. Arrow P, Brennan D, Mackean T, McPhee R, Kularatna S, Jamieson L. Evaluation of the ECOHIS and the CARIES-QC among an Australian “Aboriginal” population. Quality of Life Research. 2021;30(2):531–42.

27. Carvalho TS, Abanto J, Pinheiro ECM, Lussi A, Bönecker M. Early childhood caries and psychological perceptions on child’s oral health increase the feeling of guilt in parents: an epidemiological survey. International Journal of Paediatric Dentistry. 2018;28(1):23–32.

28. Randrianarivony J, Ravelomanantsoa JJ, Razanamihaja N. Evaluation of the reliability and validity of the Early Childhood Oral Health Impact Scale (ECOHIS) questionnaire translated into Malagasy. Health and Quality of Life Outcomes. 2020;18(1):1–11.

29. Rezaei S, Pulok MH, Moghadam TZ, Zandian H. Socioeconomic-related inequalities in dental care utilization in northwestern Iran. Clinical, Cosmetic and Investigational Dentistry. 2020;12:181–9.

30. Mizuta A, Aida J, Nakamura M, Ojima T. Does the association between guardians’ sense of coherence and their children’s untreated caries differ according to socioeconomic status? International Journal of Environmental Research and Public Health. 2020;17(5):1–11.

31. Terwee CB, Bot SDM, de Boer MR, van der Windt DAWM, Knol DL, Dekker J, et al. Quality criteria were proposed for measurement properties of health status questionnaires. Journal of Clinical Epidemiology. 2007;60(1):34–42.

